# Potential utility of plasma p-tau and NfL as surrogate biomarkers for preventive clinical trials

**DOI:** 10.1101/2022.08.17.22278853

**Authors:** Pâmela C. L Ferreira, João Pedro Ferrari-Souza, Cécile Tissot, Bruna Bellaver, Douglas T. Leffa, Firoza Z. Lussier, Guilherme Povala, Joseph Therriault, Andrea L. Benedet, Nicholas J. Ashton, Ann D. Cohen, Oscar L. Lopez, Dana L. Tudorascu, William E. Klunk, Jean-Paul Soucy, Serge Gauthier, Victor L. Villemagne, Henrik Zetterberg, Kaj Blennow, Pedro Rosa-Neto, Eduardo R Zimmer, Thomas K. Karikari, Tharick A. Pascoal, Alzheimer’s Disease Neuroimaging Initiative

## Abstract

**Background:** Although longitudinal changes in plasma phosphorylated tau 181 (p-tau181) and neurofilament light (NfL) correlate with Alzheimer’s disease (AD) progression, it is unknown whether these changes can be used to monitor drug effects in preventive clinical trials. Here, we tested the utility of changes in plasma p-tau181 and NfL as surrogate biomarkers for clinical trials focusing on cognitively unimpaired (CU) individuals.

**Methods:** We evaluated 257 CU older individuals with amyloid-beta (Aβ) positron emission tomography (PET) at baseline, as well as the baseline, up to 24-month plasma p-tau181 and NfL measures. Linear regressions and Cox-proportional hazards tested the associations of change in markers with age and clinical progression, respectively. We estimated the sample size needed to test a 25% drug effect with 80% of power at a 0.05 level on reducing changes in plasma markers.

**Results:** Longitudinal changes in plasma NfL were associated with age, while changes in plasma p-tau181 with progression to amnestic MCI. Clinical trial using p-tau181 and NfL would require 78% and 63% smaller sample sizes, respectively, for a 24-month than a 12-month follow-up. The use of Aβ positivity for enrichment had a larger impact on reducing the sample size required for trials using p-tau181 (43% reduction) than NfL (17%) as surrogate. Notably, population enrichment with intermediate levels of Aβ, rather than merely Aβ positivity, reduced the sample size by 88% for p-tau181 and 64% for NfL over 12 months, and by 73% for p-tau181 and 59% for NfL over 24 months.

**Conclusion:** Our results highlighted that changes in plasma NfL could be used as a surrogate for age-related degeneration, while longitudinal changes in plasma p-tau181 were associated with parallel clinical progression. A follow-up duration of 24 months was associated with more stable changes in plasma measures and, consequently, a greater effect size than a follow-up period of 12 months. The enrollment of CU subjects with intermediate levels of Aβ constitutes the alternative with the largest effect size for clinical trials quantifying plasma p-tau181 and NfL over 12 and 24 months.

## 1. Introduction

The preclinical stage of Alzheimer’s Disease (AD) has recently become the focus of clinical trials, based on the assumption that better therapeutic outcomes can be achieved before the onset of major neurodegeneration, Aβ and tau pathologies, and cognitive decline^1, 2^. The presence of abnormal biomarkers of Aβ plaques and tau tangles, and a lack of objective cognitive symptoms are key features of preclinical AD^3^. Even though these individuals present an elevate risk for cognitive decline, the vast majority will remain clinically stable during typical clinical trial periods of 12- to 24-month follow-up^4^. This challenges the use of changes in cognitive measures as a single primary outcome of therapeutical trials in cognitively unimpaired (CU) individuals^5^. In this context, the use of surrogate biomarkers of disease progression can be a useful addition.

Positron emission tomography (PET) and cerebrospinal fluid (CSF) biomarkers can reliably capture brain Aβ and tau pathologies, while magnetic resonance imaging (MRI) can detect neurodegenerative changes that result in atrophy. All three measures have already been proposed to monitor AD progression in clinical trials^6-9^. Recently, simpler, and more cost-effective blood-based biomarkers have been developed to estimate the presence of AD-related pathologies in the human brain^10, 11^. Plasma neurofilament light (NfL) has exhibited high sensitivity to detect neurodegeneration in AD^12-14^, and is associated with aging in CU individuals^13, 15^. Plasma phosphorylated tau (p-tau) presents both high sensitivity and specificity for detecting AD pathophysiology^16-18^ and for predicting AD-related clinical progression^16, 19-21^. Together, these results indicate that these novel blood biomarkers have the potential to be used to monitor participants’ disease progression in preventive clinical trials in CU individuals.

Although recent studies have shown that plasma biomarkers can be useful for AD clinical trials^22^, these investigations exclusively evaluated the role of plasma biomarkers in the selection of individuals that are more likely to progress over time (population enrichment). The possible utility of longitudinal changes in plasma biomarkers as a surrogate outcome measurement in CU individuals remains unexplored in the literature. Here, we tested the hypothesis that longitudinal changes in plasma NfL and p-tau181 levels can be used to monitor therapeutic response in clinical trials focusing on CU older individuals.

## 2. Materials and methods

Data used in the preparation of this article were obtained from the Alzheimer’s Disease Neuroimaging Initiative (ADNI) database (http://adni.loni.usc.edu; last accessed March 2022). Under the leadership of principal investigator Michael W. Weiner, MD, the ADNI was launched in 2003 as a public-private initiative. ADNI study was conducted according to Good Clinical Practice guidelines, US 21CFR Part 50 – Protection of Human Subjects, and Part 56 – Institutional Review Boards, and pursuant to state and federal HIPAA regulations and was approved by the Institutional Review Board of each participating site. It is a longitudinal multicenter study that has been recruiting participants from more than 50 sites across the United States and Canada. Detailed information concerning inclusion and exclusion criteria has been described previously^23^. Written informed consent for the study was obtained from all participants and/or authorized representatives. We included 257 CU older who had available data on plasma p-tau181 and NfL in at least two visits along with [^18^F]florbetapir amyloid (Aβ)-PET at the time of first plasma measurement. CU individuals had Mini-Mental State Examination (MMSE) score of 24 or higher and a clinical dementia rating (CDR) of 0. They did not have any significant neurological disease.

### 2.1 Imaging analysis

Aβ positivity was estimated using [^18^F]florbetapir PET. The standardized uptake value ratio (SUVR) images were acquired using the whole cerebellum grey matter as the reference region. A cortical composite of [^18^F]florbetapir was calculated by averaging SUVRs in the cortical gray matter of frontal, anterior and posterior cingulate, lateral parietal, and temporal regions. Aβ positivity was determined whether the composite value was greater than 1.11, based on previous validation studies^24, 25^. We used SUVR value and equations previously established by the ADNI PET core^26^ to transform the Aβ-PET SUVR to the Centiloid scale^27^. Based on ongoing clinical trials, the individuals with Centiloid values between 20-40 were classified as having intermediate Aβ levels^28^.

### 2.2 Plasma quantification

Plasma samples were collected and stored following ADNI protocols (http://adni.loni.usc.edu/methods/). The plasma p-tau181^10^ and NfL^29^ assays were measured on single molecule array (Simoa)_HD-X instruments (Quanterix, Billerica, MA, USA) at the Clinical Neurochemistry Laboratory, University of Gothenburg, Mölndal, Sweden. Baseline p-tau181 and NfL levels were considered outliers and excluded if their value was higher than three standard deviations of the population as proposed in the previous studies^19^. Based on this criterion, 10 individuals were excluded from the analyses.

### 2.3 Statistical methods

The statistical analyses were performed using R statistical software version 4.0.5 (http://www.r-project.org/). Associations between biomarkers were tested using Pearson correlation. The slope of change of plasma p-tau181 and plasma NfL levels were calculated using linear mixed-effects (LME) models with subject-specific random slopes and intercepts, as follows:

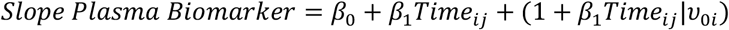

i. *β*_0_ represents the intercept;
ii. *Time*_*ij*_ represents the effect of time, fitted as a continuous measure in years from baseline;
iii. *υ*_0*i*_ is the subject-specific variation from the average intercept effect.

Time-to-event analysis was carried out to evaluate the risk of clinical progression to mild cognitive impairment (MCI). Adjusted hazard ratios (HRs) were calculated using Cox-proportional hazards models that were fitted with the following predictors: group, age, and years of education. The percentage of change in biomarkers was calculated between follow-up and baseline as follows: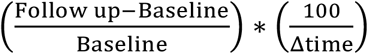. The effect size was calculated as the mean of change in biomarker in the group divided by the standard deviation^7^. We estimated the sample size required for a clinical trial testing a hypothesized 25% drug effect on longitudinal reduction in biomarker^6, 7, 30^ with 80% of power at a 5% level using a well-validated formula^7, 31^ (**Figure 1**). We used 12-month changes in tau-PET (^18^F-flortaucipir SUVR in the temporal lobe) and structural MRI (whole cortex atrophy assessed with tensor-based morphology)^7^ in Aβ positive individuals previously reported in the literature to estimate the sample size for clinical trials using neuroimaging biomarkers as surrogate outcomes.

**Figure 1.**
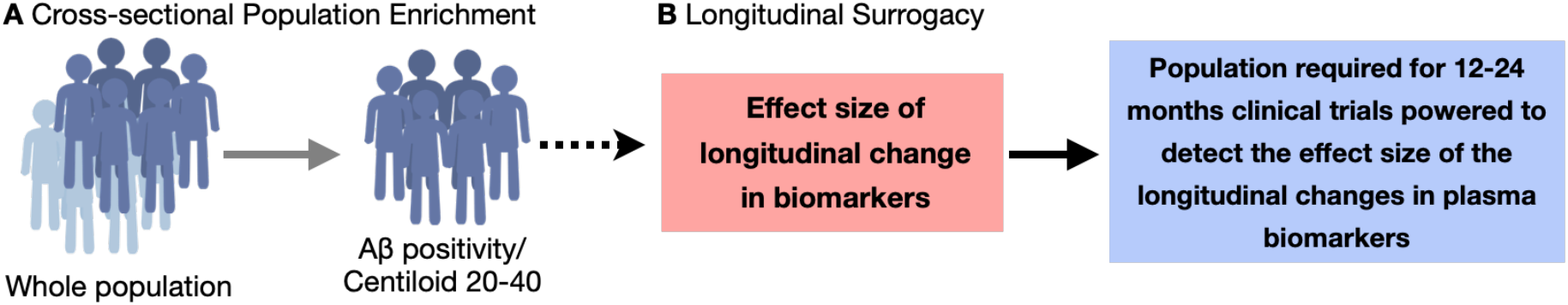
Assessment of the utility of plasma biomarker for clinical trials. (**A**) Previous studies assessed (cross-sectionally) the utility of plasma markers for selecting individuals most likely to progress over time (population enrichment). (**B**) In the present study, we evaluated the utility of plasma biomarkers to monitor drug effects (longitudinally).

## 3. Results

A total of 257 CU participants [45.5% males; mean age at baseline = 72.8 (6.16) years] were included in this study of whom 32.3% were Aβ positive. Demographics and key characteristics of the population are summarized in **Table 1** and **Supplemental Figure 1**.

**Table 1.** Participant’s demographics and key characteristics.

|  | **CU Aβ -** | **CU Aβ PET**<br>**positive** | **Intermediate**<br>**Aβ levels**<br>**(Centiloid 20-**<br>**40)** |
| --- | --- | --- | --- |
| **No.** | 174 | 83 | 25 |
| **Age at baseline, years** | 71.8 (6.02) | 74.8 (5.90) | 76.6 (6.41) |
| **Male, No. (%)** | 88 (51.2%) | 27 (32.9%) | 20 (38.5%) |
| **Education, years** | 16.9 (2.46) | 16.2 (2.74) | 16.0 (3.18) |
| ***APOE* ε4, No. (%)** | 38 (22.1%) | 35 (42.7%) | 16 (30.8%) |
| **Global Aβ PET SUVR at baseline** | 1.02 (0.05) | 1.32 (0.178) | 1.15 (0.03) |
| **Plasma p-tau181 at baseline, pg/ml** | 14.1 (10.4) | 17.1 (7.8) | 15.3 (13.2) |
| **Plasma NfL at baseline, pg/ml** | 32.4 (10.8) | 37.1 (14.0) | 36.1 (15.8) |

Continuous variables are presented as mean (SD). *APOE* $\epsilon$ 4 = Apolipoprotein E $\epsilon$ 4; p-tau181 = tau phosphorylated at threonine 181; NfL = neurofilament light.

### 3.1 Changes in plasma biomarkers as a function of their baseline levels

We observed a negative correlation between baseline plasma p-tau181 levels and its slope of change over 24 months follow-up (r = −0.17, P = 0.0012). By contrast, we found a positive association between baseline plasma NfL concentrations and plasma NfL slope of change (r = 0.59, P < 0.001) (**Supplemental Figure 2)**.

### 3.2 Association of change in plasma biomarkers with age and clinical progression

Longitudinal changes in plasma p-tau181 were not significantly correlated with age (r = 0.09, P = 0.087, **Supplemental Figure 3**), while longitudinal plasma NfL significantly correlated with participants’ age (r = 0.48, P < 0.001, **Supplemental Figure 3**). On the other hand, changes in plasma p-tau181 were significantly associated with an increased risk of clinical progression to MCI over 24 months (P = 0.037, **Figure 2**), whereas changes in plasma NfL were not (P = 0.263, **Figure 2**).

**Figure 2.**
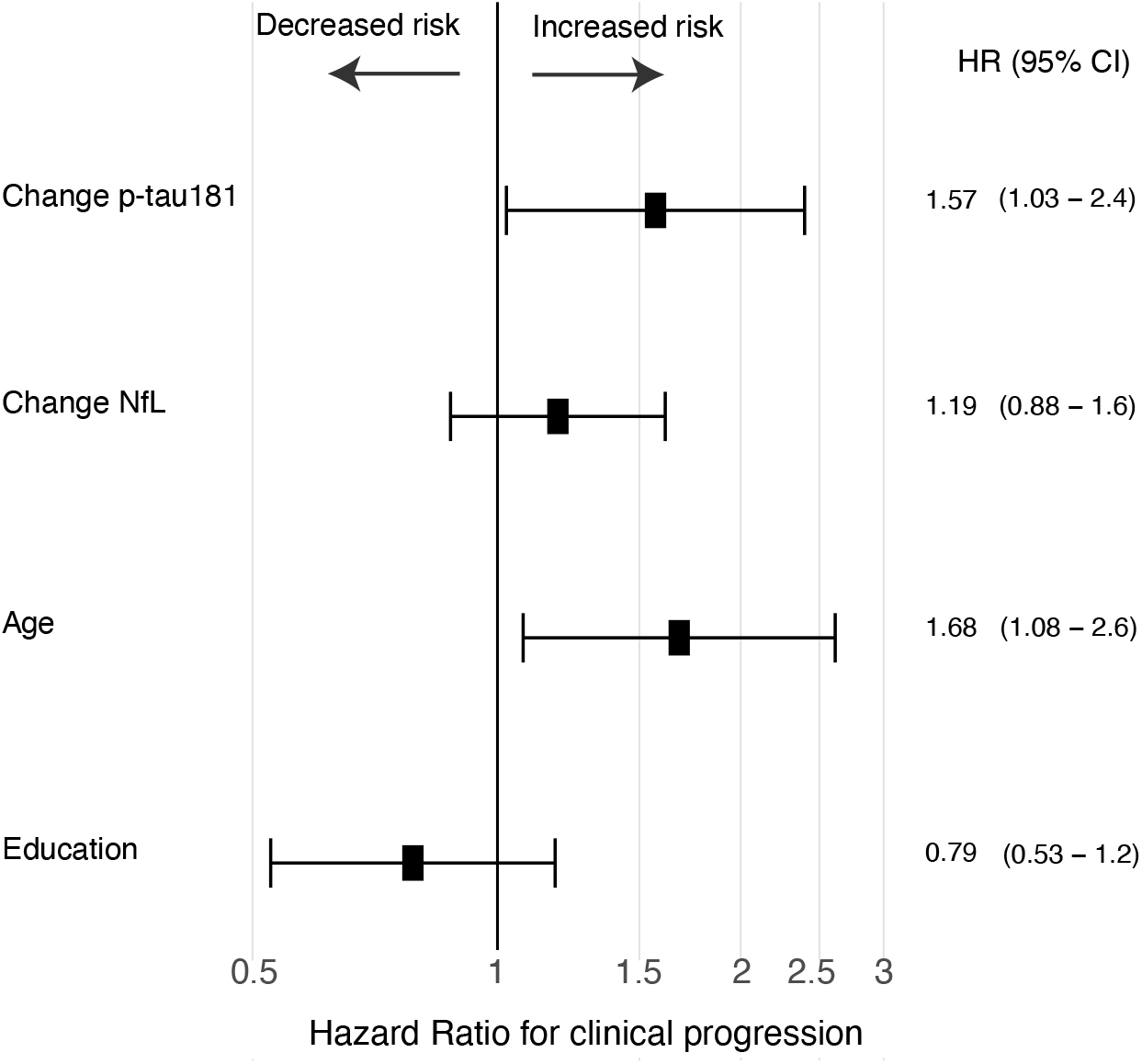
Longitudinal change in plasma p-tau181, but not in NfL, associates with an increased risk of progression to MCI. The squares represent the hazard ratio values for a clinical progression from CU to MCI over 24 months. The model was adjusted for age and years of education.

### 3.3 Longitudinal change and effect size of plasma biomarkers

The percentage of change in plasma p-tau181 and NfL (and their effect sizes) were calculated at 12- and 24-month follow-up visits in relation to their baseline values (**Figure 3A-B; Supplemental Figure 4)**. Changes in plasma p-tau181 and NfL were not significantly different from zero over 12 months in the whole population, Aβ positive, or individuals with intermediate Aβ levels. In contrast, we found a significant increase in the percentage of change of plasma p-tau181 at 24 months [CU whole population, mean: 13.84 % (95% CI, 6.4 to 21.24); Aβ negative, mean: 11.52% (95% CI, 2.8 to 20.4); Aβ positive, mean: 12.6% (95% CI, 2.2 to 23.1); intermediate Aβ ranges, mean 17.11% (95% CI, 2.9 to 31.25)]. P-tau181 effect size was numerically higher in 24 months (CU whole population: 0.23; Aβ negative: 0.21; Aβ positive: 0.34; intermediate Aβ levels: 0.44) than at 12 months (CU whole population: 0.08; Aβ negative: 0.09; Aβ positive: 0.14; intermediate Aβ levels: 0.28) for all groups. Similarly, we observed a significant increase in plasma NfL at 24 months [CU whole population, mean: 12.03 % (95% CI, 7.9 to 16.15); CU Aβ negative, mean: 11.5 % (95% CI, 6.3 to 16.3); Aβ positive, mean: 14.1% (95% CI, 6.2 to 22.1); intermediate Aβ levels, mean 19.45% (95% CI, 8.4 to 30.5)]. NfL effect size was higher at 24 months (CU whole population: 0.38; CU Aβ negative: 0.39; CU Aβ positive: 0.41; intermediate Aβ levels: 0.58) than 12 months (CU whole population: 0.24; Aβ negative: 0.28; Aβ positive: 0.25; intermediate Aβ levels: 0.39).

**Figure 3.**
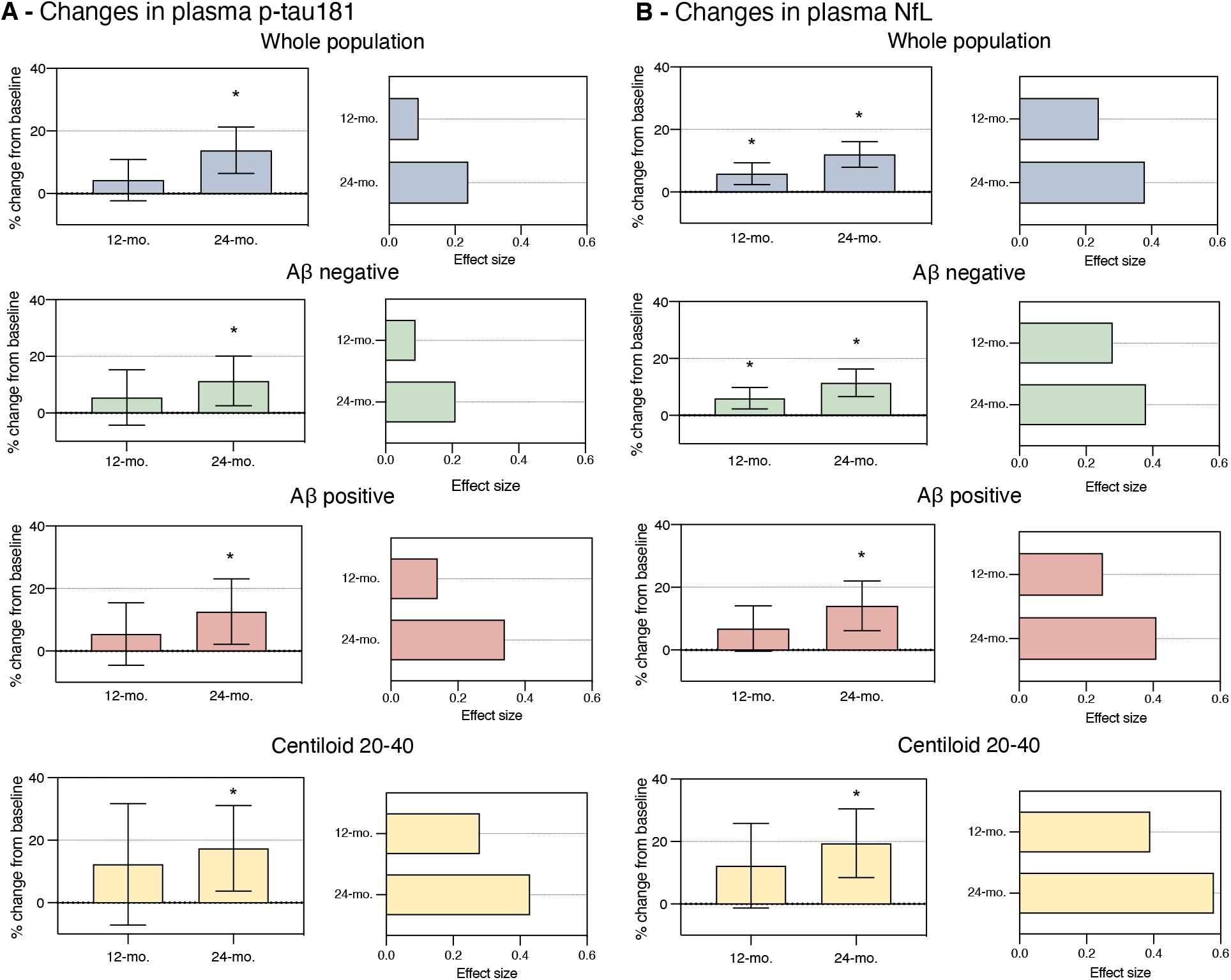
Percentage of change and effect size of plasma biomarkers over 12 and 24 months. The bar plots show the percentage of changes with their respective 95% confidence intervals for plasma (**A**) p-tau181 and (**B**) NfL concentrations in CU older individuals over 12 months and over 24 months in relation to the biomarker value at the baseline visit. The 12- and 24-month follow-ups showed a similar annualized rate of progression. The effect size at 24 months was larger due to both a greater mean of progression and a relatively more stable change among participants (smaller standard deviation (SD)). The effect size was calculated as the ratio between the mean and SD of the percentage of change overtime points. The higher the effect size, the smaller the measure’s variability, which indicates a more precise populational estimate. (*) indicates that the 95% confidence interval did not cross the zero line, and therefore, the longitudinal progression was significantly different from zero.

### 3.4 Sample size required for clinical trials in CU individuals

We estimated the sample size required for a clinical trial performed over 12- and 24-months using plasma p-tau181 or NfL as surrogate markers. Therapeutic clinical trials performed over 24 months would require 78% and 63% smaller sample sizes than 12-month clinical trials using plasma p-tau181 and NfL, respectively (**Table 2**).

**Table 2.** Sample sizes per study arm required for clinical trials enriched using different strategies.

| | All CU | CU Aβ -PET<br>positive | Intermediate<br>Aβ levels<br>(Centiloid<br>20-40) | *APOE* ε4<br>carrier | CU Aβ CSF<br>positive |
| --- | --- | --- | --- | --- | --- |
| **Plasma p-tau181** |  |  |  |  |  |
| 12 months | 28,090 | 11,796 | 3,112 | 5,974 | 6,786 |
| 24 months | 4,442 | 2,520 | 1,216 | 14,572 | 5,535 |
| **Plasma NfL** |  |  |  |  |  |
| 12 months | 4,556 | 3,886 | 1,608 | 4,540 | 4,470 |
| 24 months | 1,724 | 1,434 | 698 | 3,478 | 1,816 |

Sample size estimates per study arm for a clinical trial targeting a 25% reduction in change in plasma biomarker with 80% power and 0.05 alpha. *APOE* $\epsilon$ 4 = Apolipoprotein E $\epsilon$ 4.

A 24-month clinical trial using plasma p-tau181 as a surrogate marker, including the entire CU population, would require 4,442 individuals per study arm. In comparison, a clinical trial including only CU Aβ positive would require 2,520 individuals per study arm (reduction of 43%), and a trial with intermediate Aβ levels would require 1,216 individuals per study arm (reduction of 73%) (**Figure 4A**). A 24-month clinical trial using plasma NfL as a surrogate marker, including the entire CU population, would require 1,724 individuals per study arm. In comparison, a clinical trial including only CU Aβ positive would require 1,432 individuals per study arm (reduction of 32%), and a trial with intermediate Aβ levels would require 698 individuals per study arm (reduction of 59%) (**Figure 4A**).

**Figure 4.**
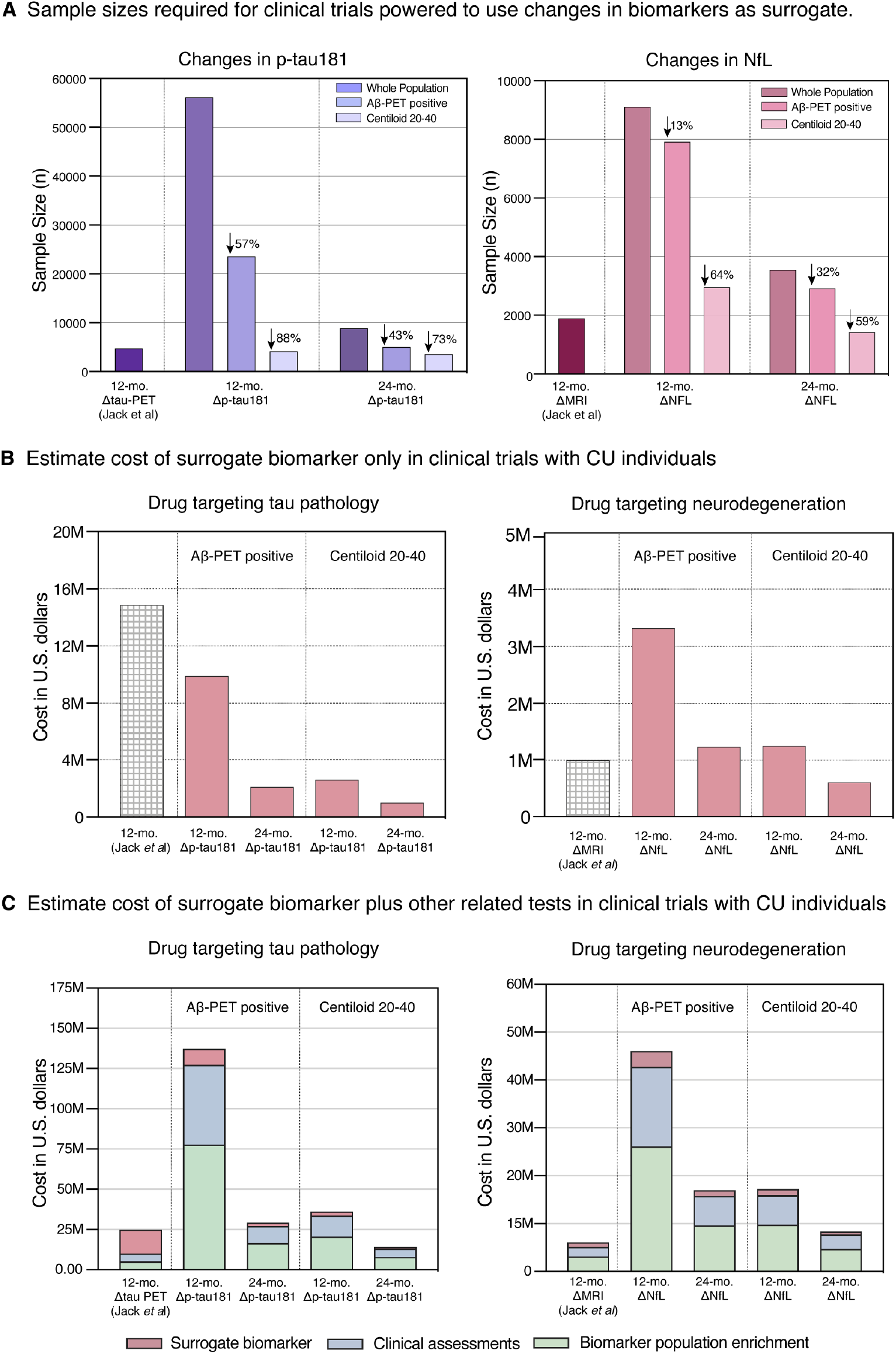
Cost-effectiveness of plasma biomarkers as surrogates for clinical trials in CU individuals. **(A)** Sample sizes required for hypothetical clinical trials powered to use plasma biomarkers to monitor drug effects in CU older whole population, Aβ positive or intermediate Aβ deposition (Centiloid 20-40). **(B)** Estimated cost with surrogate neuroimaging (Jack *et al*.^7^) and plasma (this study) biomarkers only for clinical trials powered to use changes in these biomarkers to monitor drug effects. **(C)** The estimated cost of biomarkers plus the costs with some of the other necessary tests that are influenced by total sample sizes, such as costs with the definition of Aβ positivity (using PET) for population enrichment and the standard clinical evaluations for each participant. The figure shows that using changes in plasma p-tau181 as a surrogate would reduce costs - in relation to a trial using tau-PET - if we only consider the cost with the surrogate biomarker, but not when other necessary costs that increase with sample size are considered. The costs of clinical trials using changes in tau-PET (^18^F-flortaucipir uptake in the temporal lobe) or structural MRI (tensor-based morphology cortical volume) as surrogate were estimated based on the mean and SD of a 12-month change in these biomarkers reported previously by Jack and colleagues^7^. For the calculations presented in the figure, we used the following hypothesized values: MRI = $500; PET = $3,000; plasma markers = $200; Recruitment/consenting/clinical assessment = $1,000. Assessments (excepted with the biomarker of enrichment) were calculated to 2 time-points (baseline and follow-up). We estimated an attrition rate of 10% in the calculations. Δ = longitudinal change. ↓ reduction on the sample size in relation to the whole population.

### 3.5 Cost-effectiveness analysis of plasma biomarkers for clinical trials

**Figure 4B** demonstrates that the estimated cost of a clinical trial with biomarkers only using plasma is lower than using neuroimaging biomarkers for surrogacy. **Figure 4C** shows that, in a trial including all Aβ positive individuals, the total estimated trials cost when considering surrogate biomarkers plus other related are numerically higher using plasma (more than 5-fold at 12 months and 2-fold at 24 months) than neuroimaging biomarkers for surrogacy. Interestingly, when we estimated the total cost of a trial including only individuals with intermediate Aβ levels, the cost was similar using plasma and neuroimaging biomarkers for surrogacy. Cost for clinical trials using other enrichment strategies are describe in **Supplemental Figure 4**.

### 3.6 Sample size estimation as a function of multiple drug effects

While the results mentioned above are to test a hypothesized 25% drug effect on the reduction of biomarkers increasing over time, **Figure 5** shows a progressive reduction in sample size estimates as a function of potential drug therapies with progressively higher effects on mitigating biomarkers’ changes.

**Figure 5.**
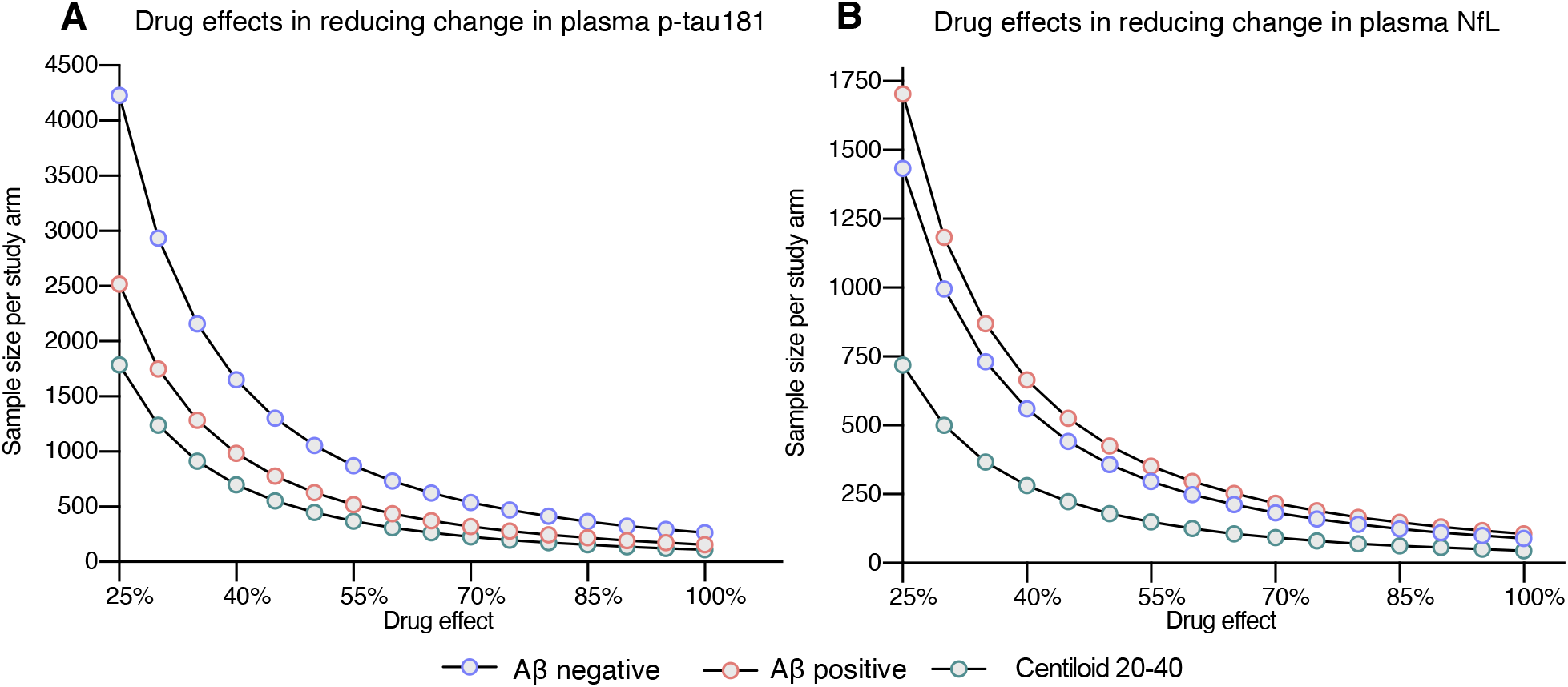
Sample sizes per study arm for a 24-month clinical trial using changes in plasma biomarkers as surrogate endpoints according to drug effects. The dots in the curves represent the sample size per study arm as a function of multiple hypothesized drug effects (greater than the tested 25%). For plasma p-tau181, a drug effect large than 60% would represent the need for a sample size of less than 500 CU Aβ positive and that have Centiloid values between 20-40 per study arm. For plasma NfL, a drug effect large than 45% would represent the need for a sample size of less than 500 CU Aβ positive and that have Centiloid values between 20-40 per study arm.

## 4. Discussion

In this study, we showed that longitudinal changes in plasma p-tau181 were associated with progression to MCI, while changes in NfL were more closely related to aging. Changes in plasma p-tau181 and NfL at 24 months, rather than 12 months, showed the potential to be used as surrogate markers in large-scale preventive clinical trials using CU older individuals. Cost-effectiveness analysis suggested that studies on CU Aβ positive will have higher total cost using plasma p-tau181 and NfL for surrogacy compared to using tau-PET and structural MRI. Interestingly, we also demonstrated that plasma studies using intermediate Aβ levels for population enrichment would be more cost-effective than using the concept of Aβ positivity.

Longitudinal changes in plasma p-tau181 were associated with clinical progression to MCI but not with age. Our results align with previous studies indicating that plasma p-tau181 predicts cognitive decline^21, 32-34^ and corroborate the notion that plasma p-tau181 can be interpreted as a relatively AD-specific biomarker even in CU individuals^21^. Given that plasma p-tau181 reflects AD-related Aβ and tau pathologies in the brain^35^, and that a large proportion of amnestic MCIs are likely to be on the AD pathway^36^. Thus, our results linking changes in p-tau181 with progression to amnestic MCI further corroborates that this marker can be used to monitor AD-related progression in CU individuals. Interestingly, we observed a negative association between baseline p-tau181 and longitudinal changes in p-tau181, suggesting that plasma p-tau181 progression decelerates in CU individuals that already present high plasma p-tau181 levels. This indicates that changes in p-tau181 are unlikely to be useful for clinical trials focused on late-stage dementia patients, where the plateau of biomarker progression has likely been reached, as was already demonstrated^17^. Together, these findings indicate that plasma p-tau181 can potentially be used for surrogacy of AD-related clinical progression in clinical trials including CU individuals.

Changes in plasma NfL were not related to progression to amnestic MCI but were highly dependent on the age of participants. These results corroborate previous studies suggesting that plasma NfL is a marker of axonal degeneration non-specific to AD-related progression, increasing as a consequence of several brain processes, including aging^13, 15, 37^. In contrast to p-tau181, changes in NfL levels were positively associated with their baseline values, suggesting that NfL is still steeply increasing in CU individuals with high plasma NfL concentrations. These results support previous literature associating plasma NfL with age-related progressive neuronal injury^37, 38^. Together, our findings suggest that plasma NfL could potentially be used as a surrogate marker of age-related neurodegeneration in clinical trials focusing on CU older individuals.

Our analysis supports that longitudinal progression in plasma p-tau181 and NfL biomarkers could potentially be used in 24-month clinical trials in CU older individuals. On the other hand, changes over 12 months showed a very high inter-subject variability and were not significantly different from zero in the CU population. We demonstrated that population enrichment with CU Aβ positive and with intermediate Aβ levels would have a higher impact in reducing the sample size needed for clinical trials using plasma p-tau181 rather than NfL as a surrogate. These results are not surprising since p-tau181 seems to be a more specific marker for AD, while NfL is highly associated with non-specific brain degeneration^13, 15, 39-41^. However, the fact that clinical trials testing 25% drug effects on biomarker reduction would require more than 5,000 and 2,800 individuals to test changes in p-tau181 and NfL, respectively, suggests that these biomarkers will be more suitable for monitoring large-scale population interventions than for formal randomized controlled trials. We used the standard 25% drug effect in our main analysis to increase comparability with several previous studies^6, 7, 30, 42^. However, it is worth mentioning that our analysis also suggested that the aforementioned scenario could be different if we consider medications with larger effect sizes. For example, a ∼50% drug effect on either p-tau181 or NfL reduction would lead to the need for a total of ∼1,000 individuals for a clinical trial using plasma biomarker as a surrogate outcome, which could be feasible for randomized controlled trials.

Surprisingly, our results suggest that using longitudinal changes in plasma p-tau181 and NfL would not reduce the cost of clinical trials using Aβ positive individuals compared to using changes in PET/MRI as a surrogate outcome. Although both tau-PET and plasma p-tau181 are postulated to reflect tau deposition in the brain^35, 43^, longitudinal changes in tau-PET reported in previous studies show more robust estimates, with less intra-subject variably in CU individuals, which translated into a considerably smaller required sample size^7^. It is known that plasma NfL and MRI are postulated to reflect non-specific neuronal damage^43^. However, the fact that structural MRI is a relatively inexpensive exam and has relatively robust longitudinal estimates corroborates that it is more cost-effective to use changes in MRI than NfL as a surrogate for neurodegeneration trials in CU individuals. While it is indisputable that blood-based biomarkers are much more accessible and less expensive than neuroimaging for a single patient, our results demonstrate that plasma p-tau181 and NfL are less cost-effective for 12- and 24-month preventive clinical trials testing a 25% drug effect in CU Aβ positive individuals due to the high variability in their longitudinal estimates and the need of larger samples sizes.

Interestingly, this study demonstrated that enrolling individuals with intermediate Aβ levels leads to smaller sample size and total cost for clinical trials using either plasma p-tau181 or NfL as a surrogate compared to studies using the concept of Aβ positivity. This was due to the fact that individuals with higher Aβ levels (centiloid > 40) showed high variability and low average of change in longitudinal plasma estimates. While some (centiloid > 40) showed elevate longitudinal changes, others plateaued their longitudinal progression. Additionally, it was already suggested that is unlikely to detect tau deposition in individuals with centiloid < 40, whereas centiloid > 40 is robustly associated with increased cortical tau deposition ^44^. Thus, the exclusion of these individuals reduces the standard deviation of changes in plasma biomarkers and consequently increases its effect size, which leads to the need for smaller sample sizes and a lower cost for clinical trials.

The strengths of this study include using data collected in multiple centers, which is more generalizable to multi-site clinical trials than using markers collected in a single site. In addition, our study included a relatively large number of participants. However, our findings must be interpreted while considering limitations. ADNI includes a selective population of highly educated, mostly white participants, generalizable to current clinical trials that have a similar population profile but do not represent a diverse general population. Thus, it would be highly desirable to reproduce our findings in a more diverse population-based cohort^45, 46^. Furthermore, the use of different blood assays for measuring the plasma biomarkers could lead to different results. Although the 25% drug effect is the standard for this type of study^6, 7, 30^, the use of different drug effects could change our conclusions. Finally, using a longer follow-up duration could lead to shifts in our results in favor of blood biomarkers, as longitudinal changes in blood markers are likely to become more meaningful over longer time frames.

## 5. Conclusion

Our results suggest that, in CU older individuals, 24-month changes in plasma p-tau181/ NfL show large inter-subject variability but could potentially be used to monitor large-scale population interventions in CU Aβ positive individuals. Clinical trials using plasma p-tau181 and NfL as surrogates with a population enriched with intermediate levels of Aβ are more cost-effective than trials using Aβ-positive individuals.

## Data Availability

All data produced in the present work are contained in the manuscript.

## Acknowledgements

Data collection and sharing for this project was funded by the Alzheimer’s Disease Neuroimaging Initiative (ADNI; National Institutes of Health Grant U01 AG024904) and DOD ADNI (Department of Defense award number W81XWH-12-2-0012). ADNI is funded by the National Institute on Aging, the National Institute of Biomedical Imaging and Bioengineering and through generous contributions from the following: AbbVie, Alzheimer’s Association; Alzheimer’s Drug Discovery Foundation; Araclon Biotech; BioClinica; Biogen; Bristol–Myers Squibb Company; CereSpir; Eisai Inc.; Elan Pharmaceuticals; Eli Lilly and Company; EuroImmun; F. Hoffmann-La Roche and its affiliated company Genentech; Fujirebio; GE Healthcare; IXICO; Janssen Alzheimer Immunotherapy Research & Development; Johnson & Johnson Pharmaceutical Research & Development; Lumosity; Lundbeck; Merck; Meso Scale Diagnostics; NeuroRx Research; Neurotrack Technologies; Novartis Pharmaceuticals Corporation; Pfizer; Piramal Imaging; Servier; Takeda Pharmaceutical Company; and Transition Therapeutics. The Canadian Institutes of Health Research is providing funds to support ADNI clinical sites in Canada. Private sector contributions are facilitated by the Foundation for the National Institutes of Health (www.fnih.org). The grantee organization is the Northern California Institute for Research and Education, and the study is coordinated by the Alzheimer’s Disease Cooperative Study at the University of California, San Diego. ADNI data are disseminated by the Laboratory for Neuro Imaging at the University of Southern California.

## References

1. Sperling RA, Rentz DM, Johnson KA, et al. The A4 study: stopping AD before symptoms begin? Sci Transl Med 2014;6:228fs213.

2. Cummings J, Lee G, Zhong K, Fonseca J, Taghva K. Alzheimer’s disease drug development pipeline: 2021. Alzheimers Dement (N Y) 2021;7:e12179.

3. Jack CR, Jr., Bennett DA, Blennow K, et al. NIA-AA Research Framework: Toward a biological definition of Alzheimer’s disease. Alzheimers Dement 2018;14:535–562.

4. Dubois B, Hampel H, Feldman HH, et al. Preclinical Alzheimer’s disease: Definition, natural history, and diagnostic criteria. Alzheimers Dement 2016;12:292–323.

5. Gauthier S, Albert M, Fox N, et al. Why has therapy development for dementia failed in the last two decades? Alzheimers Dement 2016;12:60–64.

6. Pascoal TA, Mathotaarachchi S, Shin M, et al. Amyloid and tau signatures of brain metabolic decline in preclinical Alzheimer’s disease. Eur J Nucl Med Mol Imaging 2018;45:1021–1030.

7. Jack CR, Jr., Wiste HJ, Schwarz CG, et al. Longitudinal tau PET in ageing and Alzheimer’s disease. Brain 2018;141:1517–1528.

8. Moscoso A, Karikari TK, Grothe MJ, et al. CSF biomarkers and plasma p-tau181 as predictors of longitudinal tau accumulation: Implications for clinical trial design. Alzheimers Dement 2022.

9. Budd Haeberlein S, Aisen PS, Barkhof F, et al. Two Randomized Phase 3 Studies of Aducanumab in Early Alzheimer’s Disease. J Prev Alzheimers Dis 2022;9:197–210.

10. Karikari TK, Pascoal TA, Ashton NJ, et al. Blood phosphorylated tau 181 as a biomarker for Alzheimer’s disease: a diagnostic performance and prediction modelling study using data from four prospective cohorts. Lancet Neurol 2020;19:422–433.

11. Ashton NJ, Pascoal TA, Karikari TK, et al. Plasma p-tau231: a new biomarker for incipient Alzheimer’s disease pathology. Acta Neuropathol 2021;141:709–724.

12. Santangelo R, Agosta F, Masi F, et al. Plasma neurofilament light chain levels and cognitive testing as predictors of fast progression in Alzheimer’s disease. Eur J Neurol 2021;28:2980–2988.

13. Mattsson N, Cullen NC, Andreasson U, Zetterberg H, Blennow K. Association Between Longitudinal Plasma Neurofilament Light and Neurodegeneration in Patients With Alzheimer Disease. JAMA Neurol 2019;76:791–799.

14. Mattsson N, Andreasson U, Zetterberg H, Blennow K. Association of Plasma Neurofilament Light With Neurodegeneration in Patients With Alzheimer Disease. JAMA Neurol 2017;74:557–566.

15. Ashton NJ, Janelidze S, Al Khleifat A, et al. A multicentre validation study of the diagnostic value of plasma neurofilament light. Nature Communications 2021;12:3400.

16. Janelidze S, Mattsson N, Palmqvist S, et al. Plasma P-tau181 in Alzheimer’s disease: relationship to other biomarkers, differential diagnosis, neuropathology and longitudinal progression to Alzheimer’s dementia. Nat Med 2020;26:379–386.

17. Lantero Rodriguez J, Karikari TK, Suarez-Calvet M, et al. Plasma p-tau181 accurately predicts Alzheimer’s disease pathology at least 8 years prior to post-mortem and improves the clinical characterisation of cognitive decline. Acta neuropathologica 2020;140:267–278.

18. Thijssen EH, La Joie R, Wolf A, et al. Diagnostic value of plasma phosphorylated tau181 in Alzheimer’s disease and frontotemporal lobar degeneration. Nat Med 2020;26:387–397.

19. Karikari TK, Benedet AL, Ashton NJ, et al. Diagnostic performance and prediction of clinical progression of plasma phospho-tau181 in the Alzheimer’s Disease Neuroimaging Initiative. Mol Psychiatry 2020.

20. Moscoso A. Time course of phosphorylated tau181 in blood across the Alzheimer’s disease spectrum. Brain 2020.

21. Therriault J, Benedet AL, Pascoal TA, et al. Association of plasma P-tau181 with memory decline in non-demented adults. Brain Communications 2021;3.

22. Cullen NC, Leuzy A, Janelidze S, et al. Plasma biomarkers of Alzheimer’s disease improve prediction of cognitive decline in cognitively unimpaired elderly populations. Nature Communications 2021;12:3555.

23. Petersen RC, Aisen PS, Beckett LA, et al. Alzheimer’s Disease Neuroimaging Initiative (ADNI): clinical characterization. Neurology 2010;74:201–209.

24. Landau SM, Mintun MA, Joshi AD, et al. Amyloid deposition, hypometabolism, and longitudinal cognitive decline. Ann Neurol 2012;72:578–586.

25. Landau SM, Lu M, Joshi AD, et al. Comparing positron emission tomography imaging and cerebrospinal fluid measurements of β-amyloid. Ann Neurol 2013;74:826–836.

26. Royse SK, Minhas DS, Lopresti BJ, et al. Validation of amyloid PET positivity thresholds in centiloids: a multisite PET study approach. Alzheimer’s Research & Therapy 2021;13:99.

27. Klunk WE, Koeppe RA, Price JC, et al. The Centiloid Project: standardizing quantitative amyloid plaque estimation by PET. Alzheimers Dement 2015;11:1-15.e11-14.

28. AHEAD 3-45 Study: A Study to Evaluate Efficacy and Safety of Treatment With Lecanemab in Participants With Preclinical Alzheimer’s Disease and Elevated Amyloid and Also in Participants With Early Preclinical Alzheimer’s Disease and Intermediate Amyloid. https://ClinicalTrials.gov/show/NCT04468659.

29. Gisslen M, Price RW, Andreasson U, et al. Plasma Concentration of the Neurofilament Light Protein (NFL) is a Biomarker of CNS Injury in HIV Infection: A Cross-Sectional Study. EBioMedicine 2016;3:135–140.

30. Fox NC, Cousens S, Scahill R, Harvey RJ, Rossor MN. Using serial registered brain magnetic resonance imaging to measure disease progression in Alzheimer disease: power calculations and estimates of sample size to detect treatment effects. Arch Neurol 2000;57:339–344.

31. Grill JD, D. L, Lu PH, et al. Estimating sample sizes for predementia Alzheimer’s trials based on the Alzheimer’s Disease Neuroimaging Initiative. Neurobiol Aging 2013;34:62–72.

32. Moscoso A, Grothe MJ, Ashton NJ, et al. Longitudinal Associations of Blood Phosphorylated Tau181 and Neurofilament Light Chain With Neurodegeneration in Alzheimer Disease. JAMA Neurol 2021.

33. Wang YL, Chen J, Du ZL, et al. Plasma p-tau181 Level Predicts Neurodegeneration and Progression to Alzheimer’s Dementia: A Longitudinal Study. Front Neurol 2021;12:695696.

34. Thijssen EH, La Joie R, Wolf A, et al. Diagnostic value of plasma phosphorylated tau181 in Alzheimer’s disease and frontotemporal lobar degeneration. Nat Med 2020;26:387–397.

35. Molinuevo JL, Ayton S, Batrla R, et al. Current state of Alzheimer’s fluid biomarkers. Acta Neuropathol 2018;136:821–853.

36. Langa KM, Levine DA. The diagnosis and management of mild cognitive impairment: a clinical review. Jama 2014;312:2551–2561.

37. Nyberg L, Lundquist A, Nordin Adolfsson A, et al. Elevated plasma neurofilament light in aging reflects brain white-matter alterations but does not predict cognitive decline or Alzheimer’s disease. Alzheimers Dement (Amst) 2020;12:e12050.

38. Khalil M, Pirpamer L, Hofer E, et al. Serum neurofilament light levels in normal aging and their association with morphologic brain changes. Nature Communications 2020;11:812.

39. Hampel H, O’Bryant SE, Molinuevo JL, et al. Blood-based biomarkers for Alzheimer disease: mapping the road to the clinic. Nat Rev Neurol 2018;14:639–652.

40. Hansson O, Janelidze S, Hall S, et al. Blood-based NfL: A biomarker for differential diagnosis of parkinsonian disorder. Neurology 2017;88:930–937.

41. Bridel C, van Wieringen WN, Zetterberg H, et al. Diagnostic Value of Cerebrospinal Fluid Neurofilament Light Protein in Neurology: A Systematic Review and Meta-analysis. JAMA Neurol 2019;76:1035–1048.

42. Holland D, McEvoy LK, Desikan RS, Dale AM. Enrichment and stratification for predementia Alzheimer disease clinical trials. PLoS One 2012;7:e47739.

43. Hansson O. Biomarkers for neurodegenerative diseases. Nature Medicine 2021;27:954–963.

44. Doré V, Krishnadas N, Bourgeat P, et al. Relationship between amyloid and tau levels and its impact on tau spreading. Eur J Nucl Med Mol Imaging 2021;48:2225–2232.

45. Morris JC, Schindler SE, McCue LM, et al. Assessment of Racial Disparities in Biomarkers for Alzheimer Disease. JAMA Neurol 2019;76:264–273.

46. Schindler SE, Karikari TK, Ashton NJ, et al. Effect of Race on Prediction of Brain Amyloidosis by Plasma Aβ42/Aβ40, Phosphorylated Tau, and Neurofilament Light. Neurology 2022:10.1212/WNL.0000000000200358.

