## Supplemental Table 1 for "Potential utility of plasma p-tau and NfL as surrogate biomarkers for preventive clinical trials"

### Supplementary information

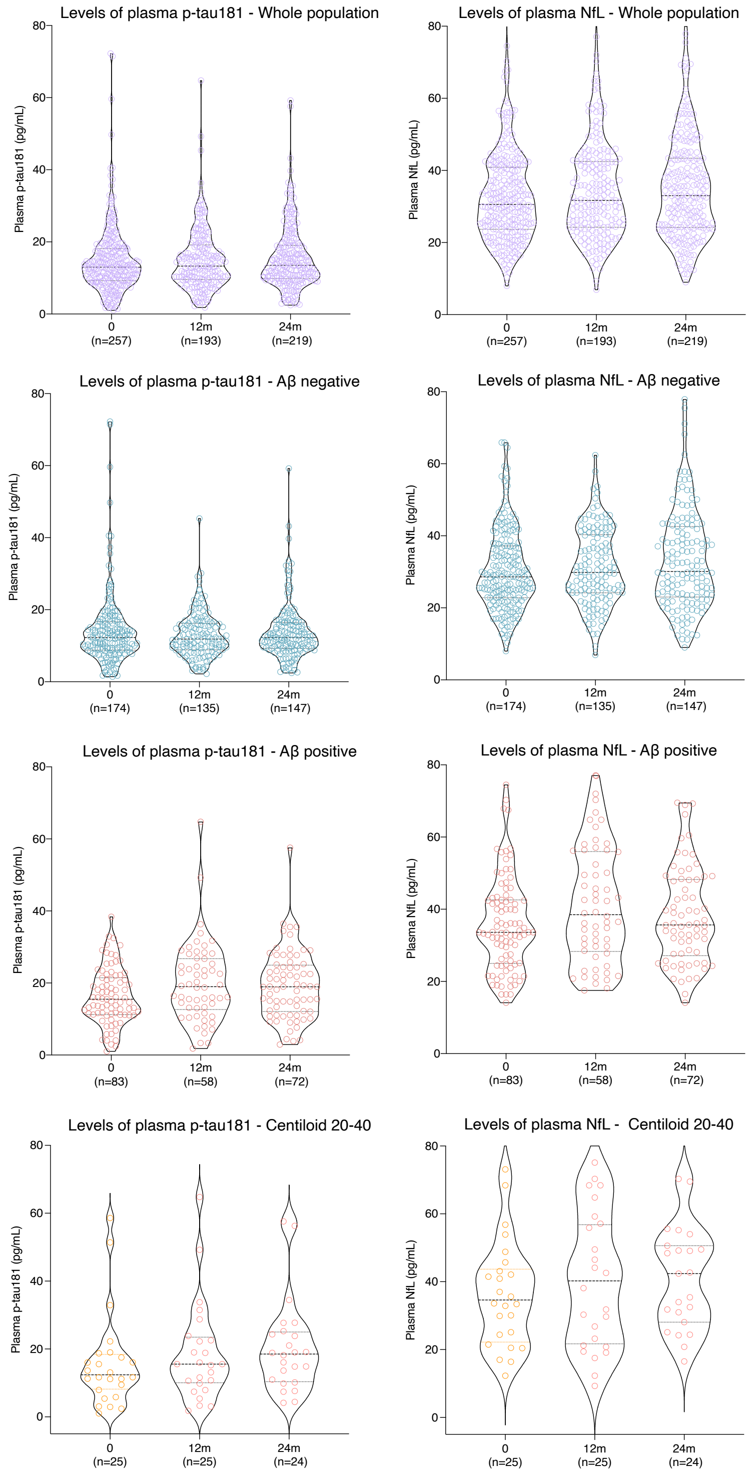


**Supplemental Figure 1. Plasma p-tau181 and NfL estimates across time frames.** The violin plots show the overall levels of plasma p-tau181and NfL concentrations across CU older individuals available in each visit and according to Aβ status. Note that violin plots at follow-up visits (12-24 months) are only partially composed by the same individuals. There are some individuals that had baseline and 12-month follow-up only, and others had baseline and 24-month follow-up only. This limits the comparison of the rates of changes between 12 and 24 months.

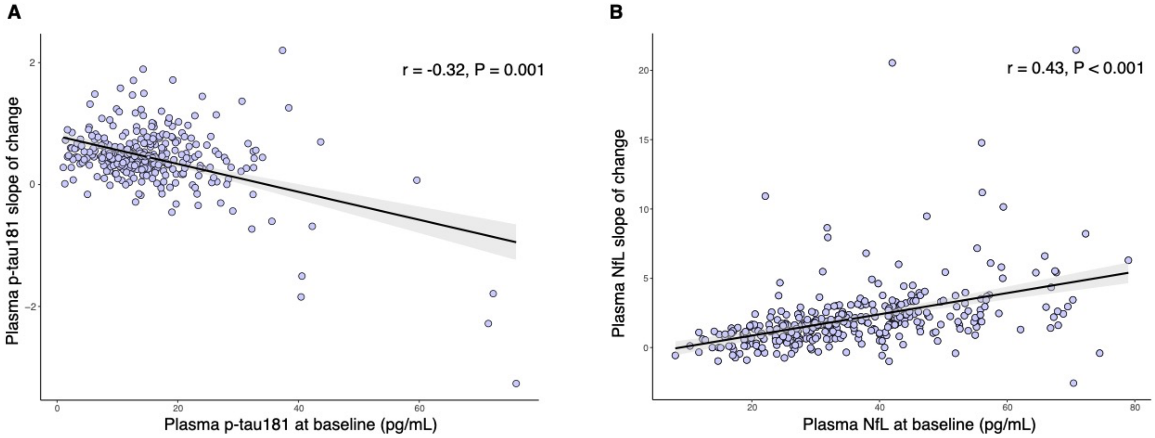


**Supplemental Figure 2. Correlation between baseline biomarkers levels and their correspondent longitudinal changes in CU individuals.** (**A**) The plot shows a negative significant Pearson correlation between the baseline and longitudinal changes in p-tau181 levels. (**B**) The plot shows a positive Pearson correlation between the baseline and longitudinal changes in NfL levels.

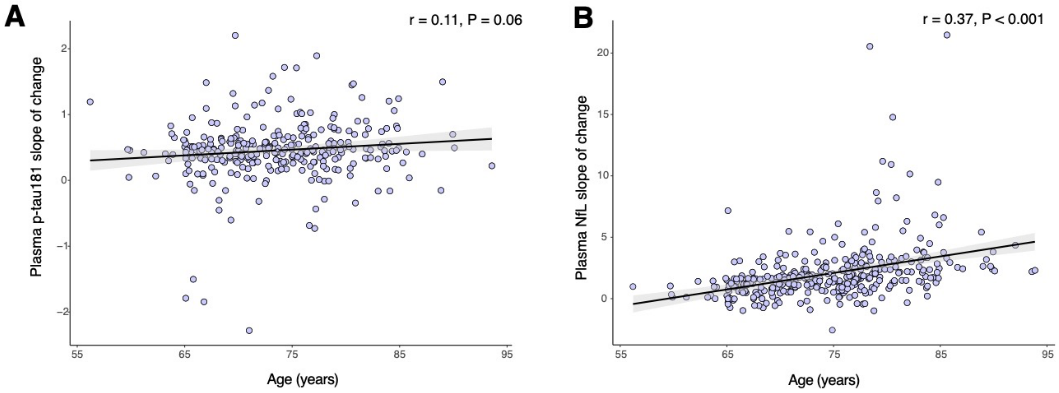


**Supplemental Figure 3 – Longitudinal change in plasma NfL, but not in p-tau181, positively associates with participants' age at baseline.** The plot (**A)** shows no significant association between longitudinal changes in p-tau181 levels and age at baseline. The plot (**B)** shows a significant association between longitudinal changes in NfL levels and age at baseline.

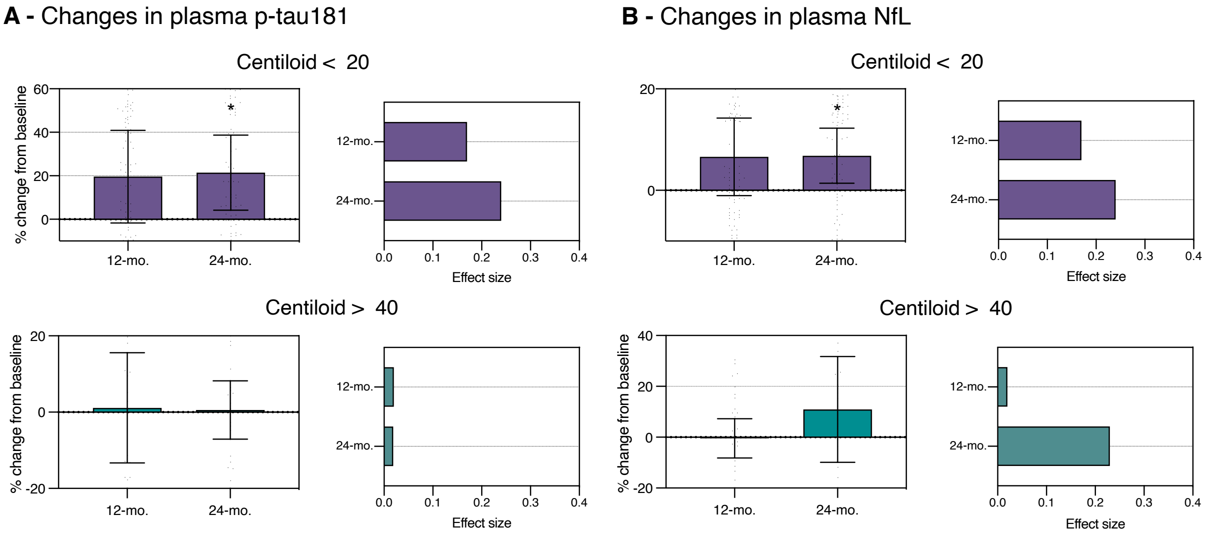


**Supplemental Figure 4. Percentage of change and effect size of plasma biomarkers over 12 and 24 months in individuals with the lowest and highest baseline Aβ.** The bar plots show the percentage of changes with their respective 95% confidence intervals for plasma (**A**) p-tau181 and (**B**) NfL concentrations in CU older individuals over 12 months and over 24 months in relation to the biomarker value at the baseline visit. The effect size was calculated as the ratio between the mean and SD of the percentage of change overtime points. (*) indicates that the 95% confidence interval did not cross the zero line, and therefore, the longitudinal progression was significantly different from zero.

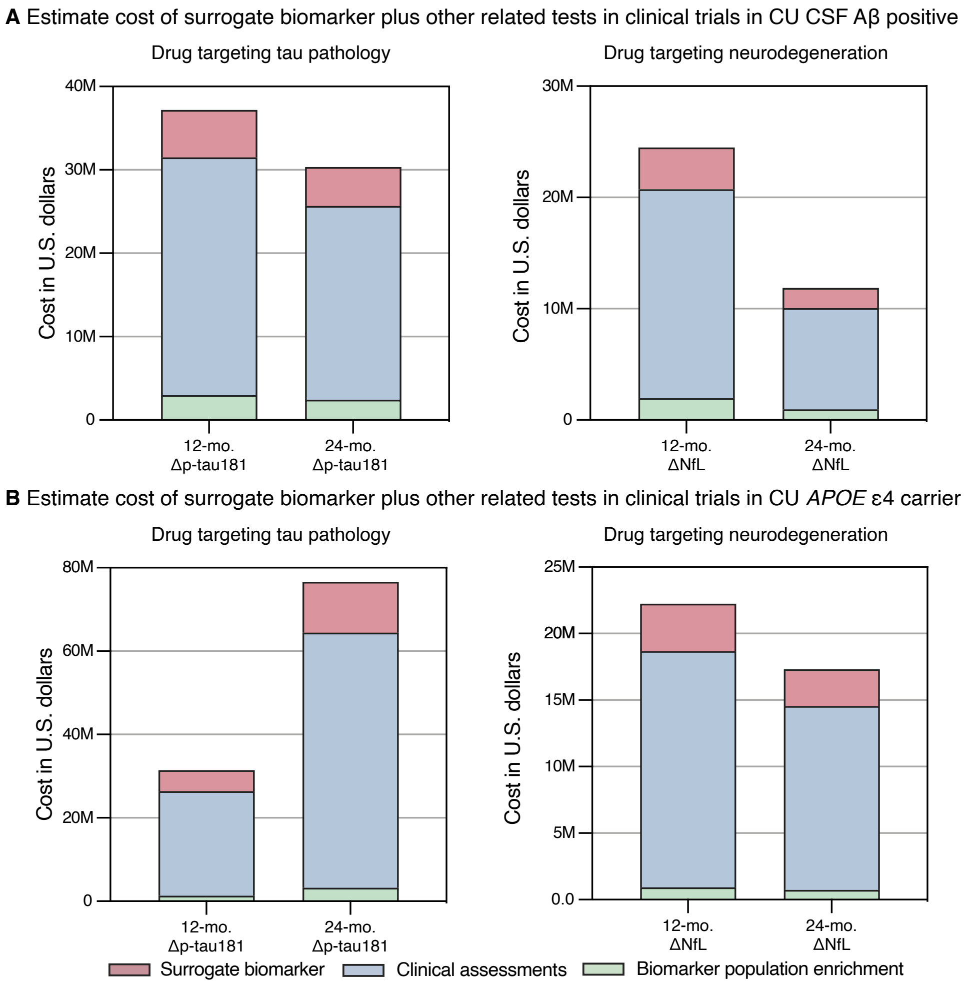


**Supplemental Figure 5. Cost-effectiveness of plasma biomarkers as surrogates for clinical trials with different population enrichment strategies.** The figure **A** shows the estimate cost of clinical trials using plasma p-tau181 and NfL as a surrogate using CSF $A\beta_{42}$ to define Aβ positivity as population enrichment strategy (Aβ positivity was defined as CSF $A\beta_{42}$ < 977 pg/mL). The figure **B** shows the estimate cost of clinical trials using *APOE* ε4 allele carriers as population enrichment strategy. For the calculations, we used the following hypothesized values: plasma markers = \$200; *APOE* ε4 allele genotype = \$100; Recruitment/consenting/clinical assessment = \$1,000. Assessments (excepted with the biomarker of enrichment) were calculated to 2 time-points (baseline and follow-up). We estimated an attrition rate of 10% in the calculations. Δ = longitudinal change.

**Supplemental Table 1: Repeated measures estimates of the slopes used in the supplementary figure 2-3.**

| Effect | β (SE) | t (DF) | p-value | 95% CI for β |
|---|---|---|---|---|
| **p-tau181** | | | | |
| Intercept | 16.79 (1.27) | 13.25 (340.6) | <0.0001 | (14.3 to 19.3) |
| Years from baseline | 0.10 (0.45) | 1.18 (238.2) | 0.817 | (-0.78 to 0.98) |
| **NfL** | | | | |
| Intercept | 37.41 (1.03) | 36.21 (362) | <0.0001 | (35.5 to 39.4) |
| Years from baseline | 2.04 (0.45) | 4.6 (192) | <0.0001 | (1.15 to 2.92) |

### Supplemental Method

**Supplemental method 1.** CSF A$\beta_{1-42}$ levels were measured using fully automated Elecsys immunoassays (Roche Diagnostics).[47, 48] Measurements outside the analytical range (< 200 pg/mL or > 1700 pg/mL for A$\beta_{1-42}$) were set to their respective technical limit. Aβ positivity was defined as CSF A$\beta_{1-42}$ < 977 pg/mL.
